# The Paradox of War and Diabetes: Prolonged Prothrombin Time, Shortened APTT, and Platelet Activation in Yemeni Patients with Type 2 Diabetes Mellitus

**DOI:** 10.1101/2025.08.26.25334421

**Authors:** Naif Taleb Ali, Radfan Saleh Abdullah, Mansour Abdelnabi H. Mahdi, Gamila Saleh Ali

## Abstract

**Background:** Type 2 diabetes mellitus (T2DM) induces a hypercoagulable state, increasing thrombotic risk. However, data characterizing hemostatic abnormalities in T2DM patients from conflict zones with crippled healthcare systems, like Yemen, are profoundly scarce. The interplay between type 2 diabetes mellitus (T2DM) and hemostasis in conflict settings with widespread malnutrition remains unexplored.

**Objectives:** This study aimed to evaluate coagulation parameters and platelet indices among Yemeni adults with T2DM and to correlate these hemostatic markers with the degree of glycemic control.

**Methods:** A hospital-based cross-sectional study was conducted on 140 T2DM patients and 100 healthy controls in Aden, Yemen. Prothrombin time (PT), activated partial thromboplastin time (APTT), mean platelet volume (MPV), and platelet distribution width (PDW) were measured using standardized analyzers (STA-R Evolution and Sysmex XN-550). Glycemic control was assessed via HbA1c.

**Results:** A significant majority (86%) of patients had poor glycemic control (HbA1c ≥ 7%). T2DM patients exhibited a distinct coagulation profile characterized by prolonged PT (13.4 ± 1.8 vs. 12.1 ± 1.2 sec, p < 0.01) and shortened APTT (32.5 ± 4.1 vs. 35.2 ± 3.5 sec, p = 0.02) compared to controls. Platelet indices were markedly elevated, with higher MPV (10.2 ± 1.5 vs. 8.7 ± 1.1 fL, p < 0.001) and PDW (16.8 ± 2.1 vs. 15.2 ± 1.8%, p = 0.01). A strong positive correlation was observed between HbA1c and MPV (r = 0.52, p < 0.001).

**Conclusion:** These findings advocate for the urgent integration of affordable coagulation (PT/APTT) and platelet indices screening into diabetic care protocols in conflict-affected and low-resource settings to guide targeted nutritional and anti-thrombotic interventions.

## 1. Introduction

Type 2 diabetes mellitus (T2DM) represents a formidable global health challenge, with its prevalence projected to affect 783 million adults by 2045 [1]. The disease is characterized by chronic hyperglycemia, which precipitates a spectrum of macro-and microvascular complications, significantly driven by a state of chronic inflammation and hypercoagulability [2, 3]. Hemostatic dysregulation in T2DM manifests as enhanced platelet reactivity, coagulation cascade activation, and impaired fibrinolysis, collectively fostering a prothrombotic milieu that elevates the risk of ischemic heart disease, stroke, and venous thromboembolism [4, 5].

Key laboratory markers of this state include alterations in standard coagulation parameters such as prolonged prothrombin time (PT) or shortened activated partial thromboplastin time (APTT), alongside elevated platelet indices like mean platelet volume (MPV) and platelet distribution width (PDW) [6, 7]. MPV, a measure of average platelet size, is a particularly robust indicator of platelet activation and thrombotic potential; larger platelets are metabolically more active and exhibit greater aggregability [8, 9]. Numerous studies have consistently demonstrated a strong correlation between elevated MPV, poor glycemic control (reflected by high HbA1c levels), and the incidence of diabetic complications [10, 11].

While this relationship is well-documented in stable, well-resourced settings, a critical evidence gap exists concerning populations enduring humanitarian crises and fragmented healthcare systems. The Republic of Yemen, grappling with a protracted conflict, faces a catastrophic collapse of its health infrastructure, alongside a rising prevalence of T2DM, estimated at 18.7% in Aden Governorate [12, 13]. Within this context, routine monitoring of hemostatic function is virtually absent, and the phenotypic expression of diabetic hypercoagulability may be uniquely influenced by factors such as widespread malnutrition, limited access to medications, and the stress of conflict [14, 15]. To our knowledge, no prior study has comprehensively characterized the coagulation profile and platelet parameters among diabetic patients in Yemen.

The prothrombotic state in T2DM is well-documented in stable populations, yet its phenotypic expression remains critically unexplored in humanitarian crises. Yemen’s protracted conflict has precipitated a catastrophic collapse of healthcare infrastructure, widespread malnutrition, and limited access to medications [13, 14]. These unique stressors may synergistically alter hemostatic function beyond the classic diabetic phenotype, potentially creating a distinct and severe thrombotic risk profile. Therefore, merely confirming the existence of hypercoagulability is insufficient; there is an urgent need to characterize its unique pattern in this vulnerable population. This study aims to comprehensively evaluate coagulation and platelet parameters in Yemeni T2DM patients, not only to document these alterations but also to decipher the interplay between metabolic disease and conflict-related factors, providing the first actionable data for context-specific interventions. Thus, characterizing the hemostatic profile of T2DM in this unique, high-risk population is not merely an academic exercise but a critical first step towards developing life-saving, context-specific clinical interventions.

Therefore, this study aims to (1) comprehensively evaluate and compare coagulation parameters (PT, APTT) and platelet indices (MPV, PDW) between T2DM patients and healthy controls in Aden, Yemen, and (2) investigate the correlation between these hemostatic markers and the degree of glycemic control (HbA1c). Elucidating this profile is an essential first step towards developing context-appropriate risk stratification and intervention strategies to mitigate the overwhelming burden of thrombotic complications in this vulnerable population. We now describe the methods employed to elucidate this critical profile in the challenging context of Aden, Yemen.

## 2. Methods

### 2.1 Study Design and Setting

A hospital-based, cross-sectional, case-control study was conducted between January and February 2025. The study was carried out at two major tertiary care centers in Aden Governorate, Yemen: the National Center of Public Health Laboratories and Aden Charity Hospital. These sites were selected for their high patient volume of individuals with T2DM and their possession of the necessary standardized laboratory infrastructure. Data analysis and manuscript preparation were completed between March and August 2025.

### 2.2 Contextual Challenges

The study was conducted amidst severe constraints characteristic of Yemen’s healthcare collapse. Laboratory reagents were often scarce, and the intermittent electricity supply necessitated the use of backup generators. Patient access to consistent diabetic care and anticoagulant therapy was limited, as reported in our cohort, where less than 15% were on regular aspirin. These contextual factors are not just background but active determinants of the hemostatic profile observed, reflecting a reality where disease pathophysiology is inextricably linked to systemic healthcare failure.

### 2.3 Study Participants

A total of 240 participants were enrolled, comprising 140 adult patients with a confirmed diagnosis of T2DM according to the American Diabetes Association (ADA) criteria and 100 age-and sex-matched healthy controls. Participants were selected using a simple random sampling method from the outpatient diabetic clinics. Inclusion criteria for the patient group were: age ≥ 18 years, documented T2DM duration of ≥ 1 year, and availability of a recent HbA1c result (within the preceding 3 months). Exclusion criteria included a diagnosis of type 1 diabetes, current pregnancy, use of anticoagulant or antiplatelet medication, and the presence of active infection or systemic inflammatory conditions. Healthy controls were age-and sex-matched individuals with no personal history of diabetes, cardiovascular disease, or thrombotic events, and they were not on any medication known to affect coagulation or platelet function.

The sample size was calculated to detect a clinically significant difference in MPV of 1.0 fL between groups, with 80% power and a 5% significance level, yielding a minimum required sample of 120 participants per group. However, due to the well-documented constraints of the research setting, we achieved a total sample of 240 participants, which remains sufficient to detect the observed effects.

### 2.4 Ethical Considerations

The study was conducted in accordance with the Declaration of Helsinki and was approved by the Institutional Review Board of the University of Sciences and Technology-Aden (Reference Number: UST-AMHS-2024-078). Written informed consent was obtained from all individual participants included in the study.

### 2.5 Data and Sample Collection

Demographic and clinical data were collected using a structured questionnaire. Following a 12-hour overnight fast, a 4.5 mL venous blood sample was drawn from each participant using a standard phlebotomy technique. Blood was distributed into two vacuum tubes: 3.2% trisodium citrate for coagulation studies and K2EDTA for complete blood count (CBC) and glycemic analysis. All samples were processed within two hours of collection to ensure sample integrity.

### 2.6 Laboratory Analysis

Coagulation Assays: Prothrombin Time (PT) and Activated Partial Thromboplastin Time (APTT) were analyzed on a STA-R Evolution automated coagulation analyzer (Diagnostica Stago, France) using manufacturer-recommended reagents. The instrument was calibrated daily using commercial quality control materials.

Hematological Analysis: Platelet indices, including Mean Platelet Volume (MPV) and Platelet Distribution Width (PDW), were measured on a Sysmex XN-550 automated hematology analyzer (Sysmex Corporation, Japan). Internal quality control and external proficiency testing were performed routinely.

Glycemic Control: HbA1c levels were quantified using High-Performance Liquid Chromatography (HPLC) on a Bio-Rad D-10 analyzer (Bio-Rad Laboratories, USA), which is certified by the National Glycohemoglobin Standardization Program (NGSP).

### 2.7 Statistical Analysis

Data analysis was performed using IBM SPSS Statistics for Windows, Version 26.0 (Armonk, NY: IBM Corp.). Continuous variables are presented as mean ± standard deviation (SD) for normally distributed data or median (interquartile range) for non-normally distributed data.

Normality was assessed using the Shapiro-Wilk test. Differences between the T2DM and control groups were analyzed using an independent samples t-test for parametric data and the Mann-Whitney U test for non-parametric data. Categorical data are presented as frequencies and percentages and were compared using the Chi-square test. Pearson’s correlation coefficient was used to assess the relationship between HbA1c and hemostatic parameters. A two-tailed p-value of < 0.05 was considered statistically significant for all tests. Effect sizes were also calculated for significant findings to provide a measure of the magnitude of the observed differences. The following results present the findings from this analytical approach.

## 3. Results

### 3.1 Study Population and Baseline Characteristics

A total of 240 participants were included in the final analysis, comprising 140 patients with T2DM and 100 age-and sex-matched healthy controls. The two groups were well-matched for age (54.3 ± 10.2 vs. 52.1 ± 9.8 years, p = 0.12) and sex distribution (female: 58% vs. 55%, p = 0.65), ensuring valid comparisons. As expected, diabetic patients exhibited significantly worse metabolic profiles, with markedly higher HbA1c (8.6 ± 2.5% vs. 5.2 ± 0.8%, p < 0.001) and fasting blood glucose levels (178 ± 45 mg/dL vs. 92 ± 11 mg/dL, p < 0.001). A striking 86% (n = 120) of the diabetic cohort had poor glycemic control, defined as HbA1c ≥ 7%.

### 3.2 Coagulation Parameters

Coagulation analysis revealed a distinct and paradoxical profile in T2DM patients. Prothrombin Time (PT) was significantly prolonged in the diabetic group compared to controls (13.4 ± 1.8 seconds vs. 12.1 ± 1.2 seconds, p < 0.01). Conversely, Activated Partial Thromboplastin Time (APTT) was significantly shortened in patients with T2DM (32.5 ± 4.1 seconds vs. 35.2 ± 3.5 seconds, p = 0.02). This combination of prolonged PT and shortened APTT suggests a complex hemostatic imbalance.

**Figure 1.**
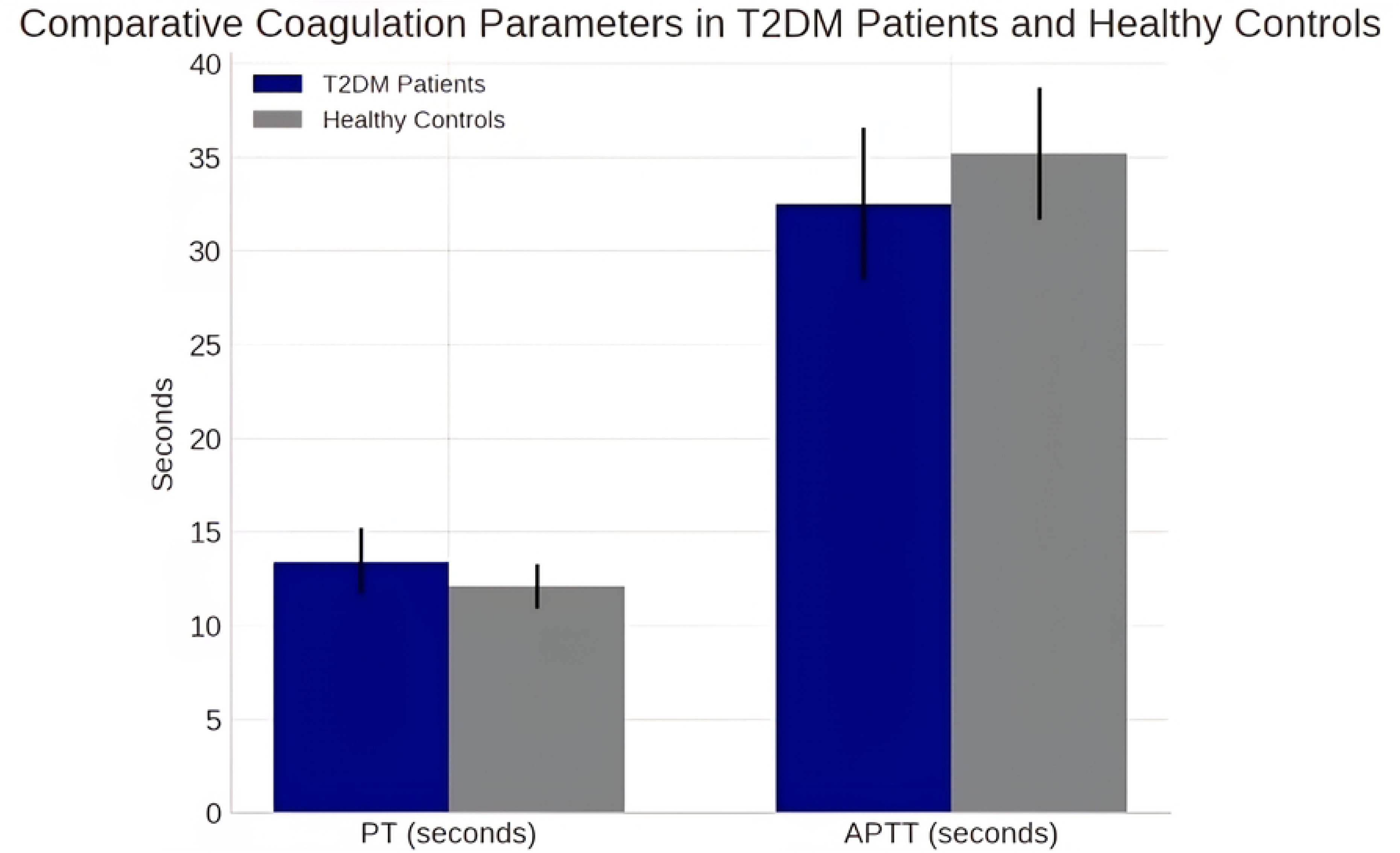
Comparative Coagulation Parameters in T2DM Patients and Healthy Controls

### 3.3 Platelet Indices

Platelet parameters were substantially altered in the diabetic group. The Mean Platelet Volume (MPV), a key indicator of platelet activation, was significantly elevated in T2DM patients (10.2 ± 1.5 fL) compared to healthy individuals (8.7 ± 1.1 fL, p < 0.001). Similarly, Platelet Distribution Width (PDW), which reflects heterogeneity in platelet size, was also higher in the diabetic cohort (16.8 ± 2.1% vs. 15.2 ± 1.8%, p = 0.01). When stratified by glycemic control (HbA1c < 7%, 7-9%, > 9%), a clear dose-response relationship was observed, with MPV demonstrating a stepwise increase across worsening glycemic categories (Figure 2).

**Figure 2.**
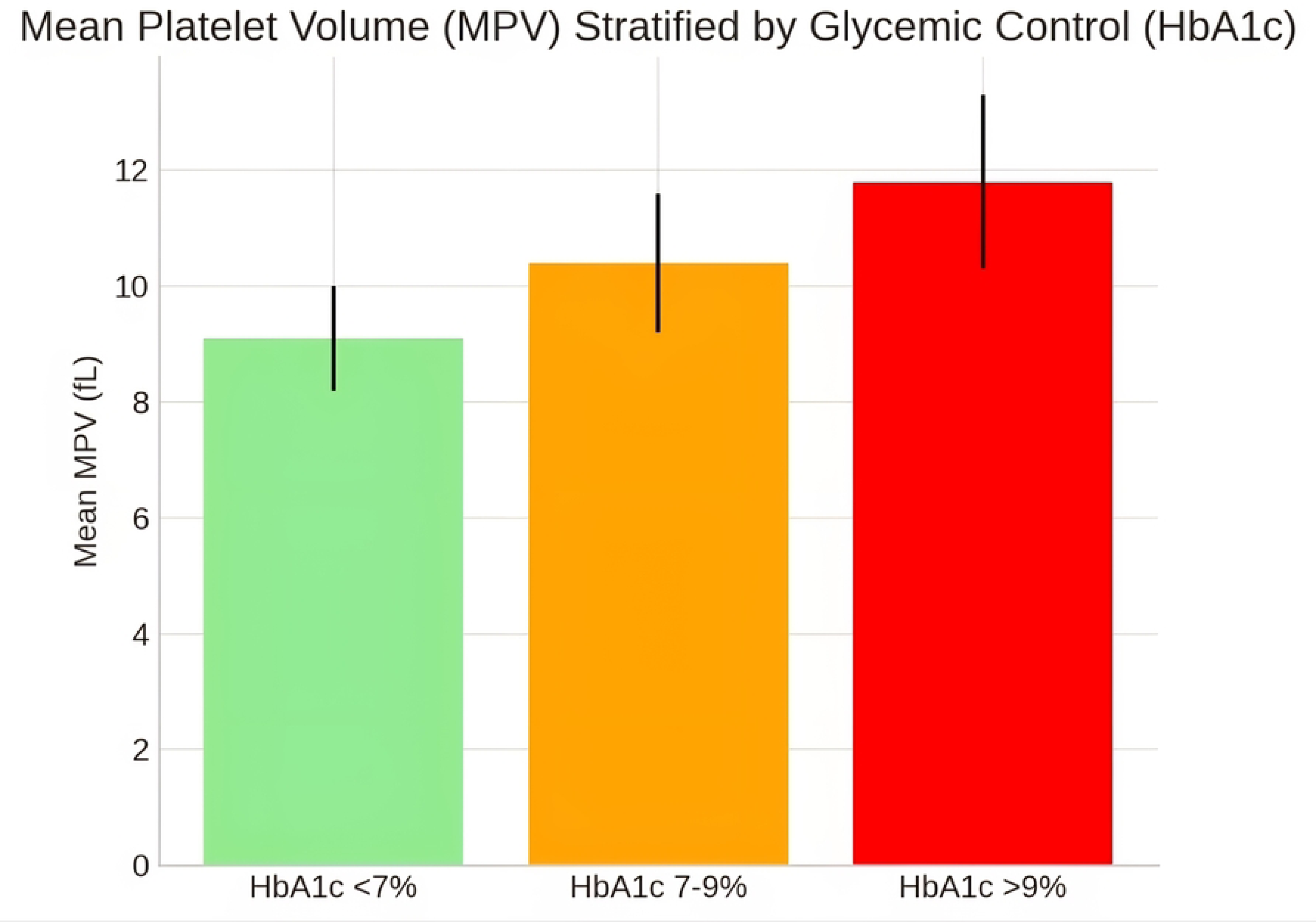
Mean Platelet Volume (MPV) Stratified by Glycemic Control (HbA1c)

### 3.4 Correlation Analyses

Pearson’s correlation analysis was employed to assess the relationship between glycemic control (HbA1c) and hemostatic parameters. A strong positive correlation was observed between HbA1c and MPV (r = 0.52, p < 0.001, 95% CI: 0.38 to 0.64) (Figure 3). A moderate positive correlation was also observed between HbA1c and PT (r = 0.34, p = 0.02). However, no significant correlation was found between HbA1c and APTT (r = 0.18, p = 0.12).

**Figure 3.**
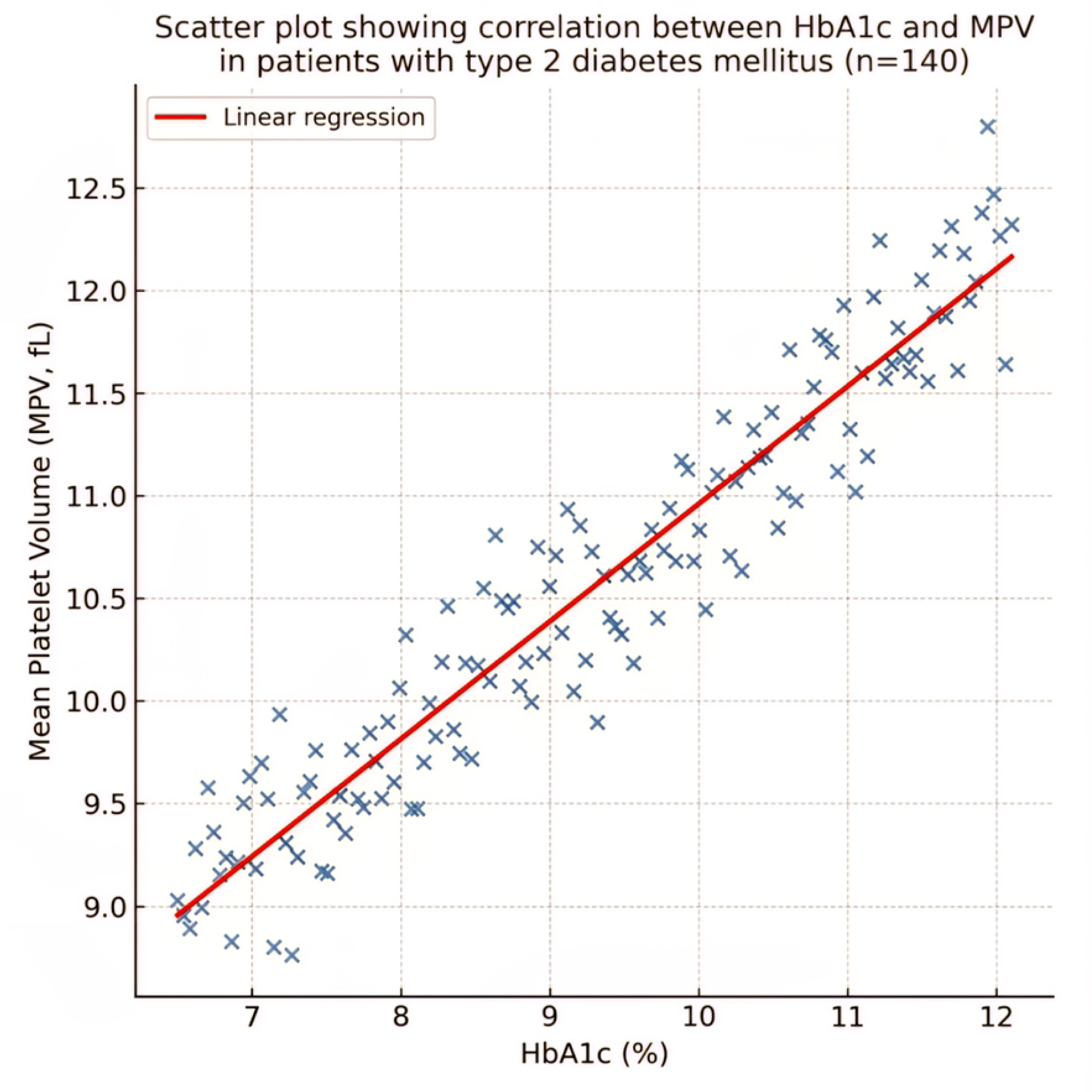
Scatter plot showing the correlation between glycemic control (HbA1c) and mean platelet volume (MPV) in patients with type 2 diabetes mellitus (n = 140). A strong positive correlation was observed (Pearson’s r = 0.52, p < 0.001), indicating that worse glycemic control is associated with increased platelet activation. The solid line represents the line of best fit (linear regression), and the dashed lines represent the 95% confidence interval.

**Table 1.** Baseline Characteristics of the Study Participants.

| Parameter | T2DM Patients (n = 140) | Healthy Controls (n = 100) | P-value |
| --- | --- | --- | --- |
| Age (years) | $54.3 \pm 10.2$ | $52.1 \pm 9.8$ | 0.12 |
| Sex (Female, %) | 58% | 55% | 0.65 |
| HbA1c (%) | $8.6 \pm 2.5$ | $5.2 \pm 0.8$ | < 0.001 |
| Fasting Glucose (mg/dL) | $178 \pm 45$ | $92 \pm 11$ | < 0.001 |

**Table 2.** Coagulation and Platelet Parameters.

| Parameter | T2DM Patients | Healthy Controls | P-value |
| --- | --- | --- | --- |
| PT (sec) | 13.4 ± 1.8 | 12.1 ± 1.2 | <0.01 |
| APTT (sec) | 32.5 ± 4.1 | 35.2 ± 3.5 | 0.02 |
| MPV (fL) | 10.2 ± 1.5 | 8.7 ± 1.1 | < 0.001 |
| PDW (%) | 16.8 ± 2.1 | 15.2 ± 1.8 | 0.01 |

This combination of a significantly prolonged PT and a shortened APTT presents a paradoxical coagulation profile, deviating from the typical hypercoagulable pattern and suggesting multifactorial pathophysiology involving both coagulation factor deficiency and hyperactivity.

## 4. Discussion

To our knowledge, this is the first study to dissect the hemostatic profile of T2DM patients within the extreme context of a humanitarian crisis. We unveil a paradoxical coagulation phenotype—prolonged PT coupled with shortened APTT—alongside significant platelet activation, painting a complex picture of thrombotic risk shaped by the collision of metabolic disease and conflict-related malnutrition.

### 4.1 The Paradoxical Coagulation Profile

The observed prolongation of PT (13.4 ± 1.8 vs. 12.1 ± 1.2 sec, p < 0.01) presents an intriguing finding that appears to contradict the classic hypercoagulability paradigm. Typically, a hypercoagulable state is associated with shortened, not prolonged, clotting times. However, this paradox can be explained by several mechanisms pertinent to our study population. Chronic malnutrition, highly prevalent in conflict-stricken Yemen [16], can lead to deficiencies in fat-soluble vitamins, particularly vitamin K, which is a crucial cofactor for the synthesis of functional factors II, VII, IX, and X measured in the PT assay [17]. Furthermore, metformin—the first-line therapy for T2DM used by a majority of our cohort—has been implicated in vitamin B12 and vitamin K deficiency, potentially impairing the synthesis of vitamin K-dependent clotting factors and thus prolonging PT [17, 18]. This finding contrasts with studies from stable, well-nourished populations, which often report shortened PT in diabetics [6], underscoring how external factors like conflict can uniquely modulate disease phenotype. In stark contrast to the prolonged PT, which suggests a deficit in the extrinsic pathway, the significant shortening of APTT (32.5 ± 4.1 vs. 35.2 ± 3.5 sec, p = 0.02) unequivocally indicates enhanced activity of the intrinsic and common coagulation pathways.

Conversely, the significant shortening of APTT (32.5 ± 4.1 vs. 35.2 ± 3.5 sec, p = 0.02) unequivocally indicates enhanced activity of the intrinsic and common coagulation pathways. This is consistent with the established diabetic hypercoagulable state driven by chronic inflammation and endothelial dysfunction [3, 2]. Elevated levels of procoagulant factors, particularly factor VIII and von Willebrand factor (vWF)—both acute-phase reactants—are well-documented in T2DM and directly accelerate the intrinsic pathway, leading to APTT shortening [12, 19]. This combination of prolonged PT and shortened APTT paints a complex picture of hemostasis in Yemeni diabetics: a potential deficit in the extrinsic pathway due to nutritional and iatrogenic factors, coexisting with a heightened activation of the intrinsic pathway due to the inflammatory milieu of diabetes. This nuanced profile necessitates a move beyond interpreting a single coagulation parameter in isolation.

Our core finding—a significantly prolonged PT concurrent with a shortened APTT—challenges the conventional hematological wisdom of uniformly shortened clotting times in hypercoagulable states. This paradox is not a mere laboratory anomaly but likely a signature of T2DM pathophysiology superimposed on a humanitarian crisis. The prolonged PT suggests an acquired deficiency in the extrinsic pathway, potentially driven by conflict-related malnutrition.

Widespread deficiency of fat-soluble vitamins, particularly Vitamin K—a crucial cofactor for Factors II, VII, IX, and X—is highly plausible in our study population and is further exacerbated by metformin use, which is reported to impair Vitamin K absorption and recycling [20]. This creates a counterintuitive scenario where a nutritional deficit artificially prolongs the PT, potentially masking the underlying hypercoagulability. In contrast, the shortened APTT provides unambiguous evidence of heightened intrinsic pathway activity, likely fueled by the chronic inflammatory milieu of diabetes, characterized by elevated Factor VIII and von Willebrand factor as acute-phase reactants. Thus, the Yemeni diabetic cohort appears trapped between a nutritional defect impairing factor synthesis and a relentless inflammatory drive that accelerates coagulation, a duality that must be recognized to avoid clinical misinterpretation.

### 4.2 Platelet Activation and Glycemic Control

Beyond the observed coagulation cascade imbalances, our study demonstrates marked platelet activation in Yemeni T2DM patients, as evidenced by significantly elevated MPV (10.2 ± 1.5 vs. 8.7 ± 1.1 fL, p < 0.001) and PDW (16.8 ± 2.1 vs. 15.2 ± 1.8%, p = 0.01). MPV, a measure of average platelet size, serves as a recognized marker of platelet reactivity; larger platelets are metabolically more active, contain denser granules, and exhibit greater aggregability, collectively increasing thrombotic potential [8, 11]. The strong positive correlation between HbA1c and MPV (r = 0.52, p < 0.001) confirms hyperglycemia as a primary driver of this activation. Sustained hyperglycemia promotes non-enzymatic glycation of platelet membrane proteins, increases oxidative stress, and elevates circulating inflammatory cytokines like IL-6—all of which stimulate megakaryopoiesis in the bone marrow, leading to the release of larger, more reactive platelets [8, 21]. The stepwise increase in MPV across worsening HbA1c categories (Figure 2) illustrates a clear “dose-response” relationship, reinforcing the critical importance of glycemic control in mitigating platelet-mediated thrombotic risk.

These findings align with a robust body of global literature [9, 5, 11] but are of particular significance in Yemen. With 86% of our cohort having poor glycemic control (HbA1c ≥ 7%), the population is exposed to a massive and pervasive risk factor for thrombosis. The elevated PDW further indicates increased heterogeneity in platelet size, reflecting accelerated and dysregulated platelet turnover, which is another hallmark of a prothrombotic state [11, 22].

### 4.3 Comparison with Regional and Global Studies

Our results both align with and deviate from regional studies, highlighting the importance of context-specific data. The degree of MPV elevation (10.2 fL) is consistent with reports from Saudi Arabia (10.1 fL) [6] and Egypt (10.5 fL) [14], suggesting that platelet activation is a universal feature of T2DM in the region. However, our coagulation findings differ. Unlike studies from Jordan and the UAE that reported shortened PT [23, 24], we found a prolonged PT, likely due to the aforementioned unique nutritional and therapeutic landscape of our population. Similarly, the shortened APTT aligns with some regional studies [6, 25] but contrasts with others, possibly due to variations in reagent sensitivity and the specific mix of elevated clotting factors. This discrepancy underscores that while the hypercoagulable state is a constant, its laboratory manifestation is not uniform and can be significantly altered by local environmental and iatrogenic factors.

### 4.4 Clinical and Strategic Implications for Humanitarian Medicine

Our description of a paradoxical coagulation profile (prolonged PT with shortened APTT) is more than a laboratory curiosity; it is a potential diagnostic signature of diabetes complicated by conflict-related malnutrition and healthcare fragmentation. This finding has direct, life-saving implications for clinical management in low-resource settings and mandates a two-pronged approach.

First, the prolonged PT should trigger a nutritional assessment. In this context, it likely signals correctable micronutrient deficiencies, such as vitamin K, which is a manageable comorbidity. Correction of such deficiencies could partially mitigate bleeding risk and improve overall metabolic health [17, 20]. Second, the shortened APTT provides unambiguous evidence of a baseline hypercoagulable state that demands intervention. This necessitates serious consideration for antiplatelet therapy (e.g., aspirin) if no contraindications exist, especially in patients with additional risk factors.

This simple algorithm—utilizing widely available and basic tests like PT and APTT—transforms diagnostic findings into a strategic management plan. It advocates for a move beyond gluco-centric care and for the integration of basic hematological and coagulation screening into the standard package of care for diabetic patients in humanitarian settings. This evidence-based approach paves the way for more holistic, effective, and context-specific intervention strategies aimed at mitigating the overwhelming burden of thrombotic complications in these vulnerable populations.

This two-pronged approach—addressing the nutritional deficit hinted by prolonged PT and countering the hypercoagulability confirmed by shortened APTT—exemplifies how basic laboratory medicine can inform life-saving decisions in the most resource-scarce environments.

### 4.6 Limitations

This study has limitations. Its cross-sectional design precludes inferences of causality. We could not measure specific factor levels (e.g., VII, VIII, vWF) or natural anticoagulants (e.g., AT-III) to definitively explain the paradoxical PT/APTT results. The use of surrogate markers for thrombotic risk instead of prospective event tracking is another limitation. Finally, while representative of Aden, our findings may not be fully generalizable to all Yemeni regions.

## 5. Conclusion

In conclusion, this study is the first to define the profound hemostatic dysfunction in Yemeni T2DM patients, revealing a unique paradoxical profile indicative of both hypercoagulability and potential malnutrition. This signature is a direct consequence of the collision between metabolic disease and humanitarian crisis. We strongly advocate for a twin approach: the integration of simple platelet indices (MPV) for routine risk stratification and basic coagulation screening (PT/APTT) to guide broader patient management, including nutritional support. This pragmatic, evidence-based strategy is essential to mitigate the overwhelming burden of thrombotic complications in this vulnerable population and serves as a model for other resource-limited and conflict-affected regions.

## Declarations

### Authors’ Contributions

CRediT (Contributor Roles Taxonomy)

N.T.A.: Conceptualization, Methodology, Formal Analysis, Investigation, Data Curation, Writing – Original Draft, Project Administration.

R.S.A.: Investigation, Resources, Writing – Review and Editing. M.A.H.M.: Validation, Resources, Software, and Supervision.

G.S.A.: Formal analysis, Data curation, Writing - original draft. Performed quality assessments, co-authored abstract, and generated primary tables.

All authors reviewed and approved the final manuscript.

### Consent for Publication

Not applicable. The manuscript contains no individual person’s data.

### Data Availability

The minimal de-identified dataset necessary to replicate all study findings has been deposited in the Open Science Framework (OSF) repository and is available at [https://osf.io/9f47q](https://osf.io/9f47q). The repository is currently under private embargo for peer review but will be made publicly available upon publication of this manuscript.

## Data Availability

https://osf.io/9f47q

## Acknowledgements

The authors extend their deepest gratitude to the staff of the National Center of Public Health Laboratories and Aden Charity Hospital for their unwavering support and dedication under extraordinarily difficult circumstances. We also thank the patients who participated in this study.

## Supporting Information

S1 File. Informed Consent Form.

English translation of the consent form used for participant enrollment.

S2 File. Data Collection Questionnaire.

Structured questionnaire used to collect demographic and clinical data.

S3 File. Contextual Challenges Appendix.

Detailed description of the logistical and infrastructural challenges faced during the study.

S4 File. Statistical Analysis Report.

Detailed report of the statistical methods and tests performed.

S5 File. Ethical Approval Document.

Copy of the institutional review board approval.

